# Acceptability of extending HPV-based cervical screening intervals from 3 to 5 years: An interview study with women in England

**DOI:** 10.1101/2022.01.12.22269119

**Authors:** Martin Nemec, Jo Waller, Jessica Barnes, Laura A.V Marlow

**Author notes:** corresponding author at: Address: Room GH0603334, 3^rd^ Floor, Research Oncology, Bermondsey Wing, Guy’s Hospital, Great Maze Pond, London SE1 9RT, UK.

## Abstract

**Objectives:** The introduction of primary HPV testing in the NHS Cervical Screening Programme in England means the screening interval for 25-49-year-olds can be extended from 3 to 5 years. We explored women’s responses to the proposed interval extension.

**Methods:** We conducted semi-structured phone/video interviews with 22 women aged 25-49 years. Participants were selected to vary in age, socioeconomics, and screening history. We explored attitudes to the current 3-year interval, then acceptability of a 5-year interval. Interviews were transcribed verbatim and analysed using Framework Analysis.

**Results:** Attitudes to the current 3-year interval varied; some wanted more frequent screening, believing cancer develops quickly. Some participants worried about the proposed change; others trusted it was evidence-based. Frequent questions concerned the rationale and safety of longer intervals, speed of cancer development, the possibility of HPV being missed or cell changes occurring between screens. Many participants felt reassured when the interval change was explained alongside the move to HPV primary screening, of which most had previously been unaware.

**Conclusions:** Communication of the interval change should be done in the context of broader information about HPV primary screening, emphasising that people who test negative for HPV are at lower risk of cell changes so can safely be screened every 5 years. The long time needed for HPV to develop into cervical cancer provides reassurance about safety, but it is important to be transparent that no screening test is perfect.

## INTRODUCTION

By the end of 2019, HPV-based screening had been introduced into the NHS Cervical Screening Programme across England.(1) Tests for high-risk HPV have higher sensitivity than cytology-based screening, significantly reducing the likelihood of false negatives.(2) The superior negative predictive value of HPV-based screening (almost 100%) (2) means that the screening interval for women aged 25-49 years can be extended from 3 to 5 years in England.(1) Other countries have recently implemented similar changes prolonging the interval between cervical screens to 5-years (e.g. Australia)(3) or 10-years (the Netherlands). (4)

Acceptability is an important consideration ahead of changes to health policy.(5) Following a recent systematic review,(5) it has been argued that “All components of the screening program should be clinically, socially and ethically acceptable to screening participants” (p. 427). However, previous research suggests that the transition to longer screening intervals may not be considered acceptable to all women.(6-8) Following changes to the cervical screening programme in Australia (the ‘renewal’, where HPV-based testing, changes to age-based eligibility and extended intervals between screens were introduced together),(9) research revealed public concerns. These included beliefs that women’s health was being endangered and devalued as a result of budget cuts, and that changes would lead to missed or late diagnoses of cervical cancer.(6) These concerns were related to limited understanding of the rationale for the interval changes as well as limited knowledge of HPV regression and the slow progression from HPV infection to cervical cancer.(10) More recently, a qualitative study suggested that Australian women wanted to be consulted about any further changes to the cervical screening programme prior to implementation.(11) Previous UK-based research suggests acceptability of extended screening intervals may be influenced by how the change is presented. A clearer presentation of the rationale for extended intervals could increase acceptability.(12)

While the term ‘acceptability’ has been used in health services research for many years,(13) it has recently been defined and operationalised as a “multi-faceted construct which reflects the extent to which people delivering or receiving a healthcare intervention consider it to be appropriate, based on anticipated or experienced cognitive and emotional responses.” (Theoretical Framework of Acceptability).(13) Sekhon et al.(14) argue that acceptability includes a number of constructs including Affective Attitude (how the person feels about the relevant healthcare intervention), Burden (how difficult participating in the intervention seems), Ethicality (how the intervention fits with one’s values), Intervention Coherence (how much the person understands the intervention), Opportunity Costs (what must be given up to engage in the intervention), Perceived Effectiveness (the perceived likelihood of the intervention achieving its purpose), and Self-efficacy (perceived ability to perform the behaviour associated with the intervention).

This study aimed to explore acceptability of the change to 5-yearly cervical screening intervals for women aged 25-49 years, ahead of implementation, drawing on the TFA as a broad framework.

## METHODS

### Design

This was an exploratory qualitative study, using one-to-one semi-structured interviews.

### Ethics Approval

The study was approved by the King’s College London ethics committee (LRS-19/20-19298 and MOD-20/21-19298) and reported using the Standards for Reporting Qualitative Research framework. (15)

### Participants

Participants were identified and contacted via email by a research recruitment agency with a large participant panel. The number of participants approached was not recorded. Those who were interested in taking part completed a short questionnaire providing information on demographic characteristics and cervical screening history. We used purposive sampling to ensure variation with respect to age (within the eligible age range: 25-49 years), socio-economic background and cervical screening experience. Participants were emailed a participant information sheet and consent form ahead of the interview. Signed or initialled consent forms were returned by email or, where this was not possible, consent was collected verbally prior to interview. Twenty-two participants each took part in a single interview and were paid £50 to thank them for their time. There were no withdrawals following consent.

### Procedure

Interviews were conducted by phone (n=12) or video call (using Microsoft Teams; n=10) according to participant preference by either JB or LM, both female post-doctoral researchers with training and experience in semi-structured interviewing and no prior relationship with the participants. Interviewers introduced themselves as researchers from King’s College London, funded by PHE. Interviews lasted around 40 minutes (mean=39 minutes, range 14 to 60 minutes) and were audio-recorded. Field notes were not used.

A topic guide was used to ensure key topics of interest were covered. The topic guide (see Supplementary File 2) included three sections: 1) views about the current screening interval, 2) views about the possibility of extended intervals, 3) reactions to key messages about the interval change. Prompts were developed for the first two sections to explore the different ways in which participants may find screening intervals acceptable or unacceptable. These were designed to encourage discussion of different elements of acceptability as identified in the Theoretical Framework of Acceptability.(14) In section 2, participants were provided with additional information explaining the rationale for the 5-year interval in a stepped way before discussing views in further detail. This was provided verbally using a script (see Topic Guide, Supplementary File 2). In section 3, participants were presented with additional messages designed to reassure women about the interval change (Supplementary File 2). This part of the interview was designed to help us develop key messages for a future study (described elsewhere), but also prompted further discussion so this part of the data was also analysed. The topic guide was not pilot tested.

### Patient and public involvement

Neither patients nor the public were involved in the design, conduct, analysis or interpretation of this study.

### Analysis

Interviews were transcribed verbatim by an external agency. Due to time-constraints, transcripts were not returned to participants for comment or correction. Analysis began after completion of the first five interviews so that initial findings could feed into an iterated topic guide when appropriate and to confirm data saturation.

We used Framework Analysis,(16) following the stages outlined by Gale et al.(17) Inductive line by line coding was performed on the first three transcripts by MN (male research assistant with experience and training in qualitative analysis and no prior relationship with the participants) and LM after which, codes were agreed for subsequent analysis and were grouped into categories using MIRO, an online platform that allows creation and movement of virtual post-it-notes. This working analytical framework was then applied to the rest of the transcripts and refined throughout the process. Transcripts were coded in order and regular discussions on modifications of the framework were held (LM and MN). After the first nine transcripts had been coded, a final analytical framework was confirmed. This framework was then used to code the remaining transcripts (MN) before charting (entering summarised data) into a framework matrix (a code by case spreadsheet) in Excel. Data were then charted (by MN and checked by LM). MN and LM then interpreted the data with insights from JW (female post-doctoral researcher with training and experience in qualitative research and no prior relationship with the participants) and JB. Participant checking was not part of our analysis plan.

## RESULTS

### Sample characteristics

We carried out 22 interviews before reaching data saturation. We used a stopping criterion of three meaning that data collection stopped at the point where no new themes had emerged in the previous three interviews.(18)

Demographic characteristics are shown in Table 1. Four women were aged 25-29 years, 14 were aged 30-39 years, and four were 40-49 years old (age range: 25-49 years, mean age = 34.9 years, SD = 6.4 years). Participants were evenly distributed across occupational social grade and white vs. ethnic minority groups. Seventeen participants reported having been screened in the last three years (up-to-date with screening) and five were overdue.

**Table 1.** Sociodemographic characteristics of the sample (n=22)

|  | N |
| --- | --- |
| Age |  |
| 25-29 years | 4 |
| 30-39 years | 14 |
| 40-49 years | 4 |
| Occupational social grade |  |
| AB (high) | 6 |
| C1 | 5 |
| C2 | 6 |
| DE (low) | 5 |
| Marital status |  |
| Single | 9 |
| Married/civil partnership/cohabiting/engaged | 10 |
| Divorced | 3 |
| Working status |  |
| Working (full-time or part-time) | 16 |
| Not working | 4 |
| Furloughed due to COVID-19 | 2 |
| Ethnicity |  |
| White | 11 |
| Mixed | 3 |
| Asian | 5 |
| Black | 3 |
| Sexual orientation |  |
| Heterosexual/straight | 16 |
| Gay/Lesbian/Bisexual | 5 |
| Prefer not to say | 1 |
| Educational level |  |
| None/GCSE or equivalent | 9 |
| A Level or equivalent | 6 |
| Postgraduate diploma | 1 |
| University degree or higher | 6 |
| Last cervical screen |  |
| Within the last 3 years | 17 |
| More than 3 years ago | 5 |
| Had the HPV vaccine? |  |
| Yes, I have | 6 |
| No/don't know | 16 |
| Heard of HPV before today? |  |
| Yes, I have heard of it | 18 |
| No, I have not heard of it | 4 |
| Self-rated HPV knowledge |  |
| Good/very good | 6 |
| Fair/poor/very poor | 16 |

### Summary of themes and processes that arose from the interviews

There were two broad primary topics within the interviews i) women’s acceptance of existing screening intervals and ii) their acceptance of the proposed 5-yearly screening interval (see Figure 1, Supplementary File 3). Within these topics specific themes related to beliefs about the existing and extended screening intervals. Throughout the interviews, as more information was provided, women’s understanding of the rationale for the change improved. Two broader influencing factors (‘personal risk’ and ‘trust and empowerment’) were also identified. Each theme is described below, with example quotes (see Appendix 1, Supplementary File 1, for additional participant quotes).

**Figure 1:**
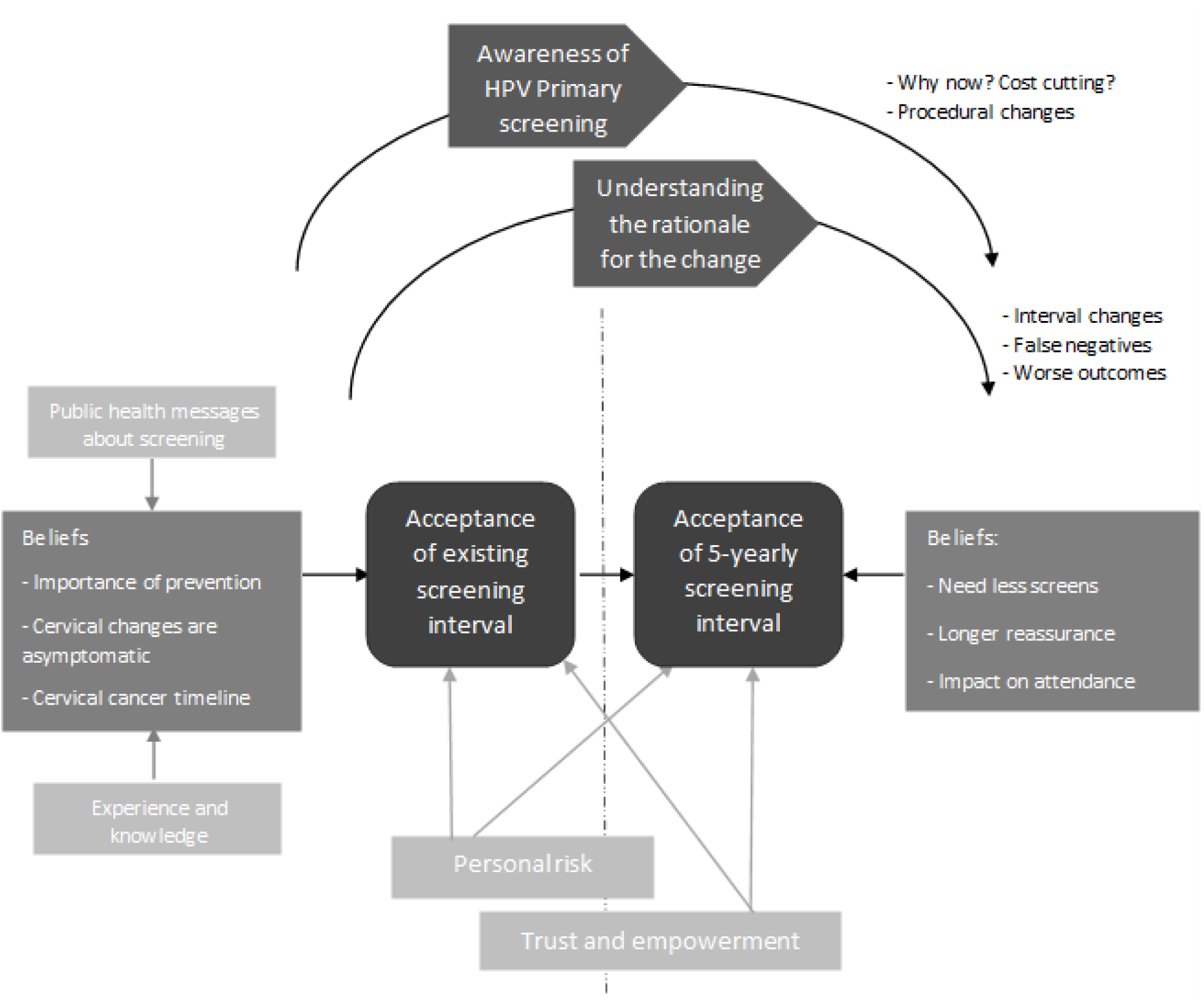
Schematic representing the themes/processes from the interviews.

### Acceptance of existing screening intervals

Acceptance of existing screening intervals was influenced by beliefs about the importance of prevention and early detection, the asymptomatic nature of cervical abnormalities and the speed of development of cervical cancer. These beliefs were driven by existing knowledge of cancer and how screening works and perceptions of existing public health messages about screening.

Regular cervical screening was felt to bring the benefits of prevention and early detection:

> *‘It may be treatable, but it might not be. And the treatability that might be linked in some way to an early detection.* …*I think in a lot of people’s mind, including mine, is that that you need to get in there quickly and you’ve got a better chance if things are treated early.’* [P13; 40-49 years; up-to-date with screening]

Cervical cancer was thought to be asymptomatic or to have symptoms only at a later stage so frequent screening was seen as way of improving outcomes. For some this meant screening more frequently than every 3 years would be considered an improvement to services:

> ‘…*without the test you wouldn’t know if you’ve got a problem with your cervix. It’s not like you’ve got a symptom and you know you’ve got a symptom*.’ [P16; 30-39 years; up-to-date]

Acceptance of screening intervals was also influenced by beliefs about the timeline of cervical cancer development. Understanding of the speed that cervical cancer develops varied:

> *‘I know that it’s supposedly, like, if you do have cervical cancer then it’s a very slow process from pre-cancerous to actually cancerous.’* [P18; 30-39 years; up-to-date]
>
> *‘…probably 36 months is a long time if, if after one smear test somebody has started developing a problem and then after 36 months compared to the last one, it might have gone if it, you know, it might go out of control*…*’ [P14; 30-39 years; overdue]*

### Acceptance of 5-yearly intervals

Initially, discussion about acceptance of 5-yearly screening reflected how women felt about the existing interval. Women who felt strongly that 3-yearly screening was important or not enough, were resistant to a 5-yearly interval, expressing shock and concern. Initial reactions centred around how a 5-yearly interval contradicted their existing beliefs about the importance of screening and early detection:

> *‘It’s just-- The whole-- Every three years has always been for prevention, to make sure you’re healthy, and now suddenly, it’s every five years. I don’t know.’* [P7; 30-39 years; up-to-date]
>
> *‘After that [Jade Goody’s death from cervical cancer] there was such a massive drive and a massive like-- Please go and get tested. Please do this. Please do that [*…*] I really felt like it made a difference. But now, if the NHS is saying, you know, you can’t even have one when people want it. That, to me, makes no sense.’* [P2; 25-29 years; up-to-date]
>
> *And I just feel like that gap between the screenings, I think personally, is much too long to catch something early enough…So it seems like quite a long time between them, yeah…I think it probably should be shorter…And then what do you do? Like how do you-- Because you’re not getting the screening, how do you know that it hasn’t progressed to abnormal cells or to cervical cancer within that five-year period if you’re not being screened?’* [P6; 25-29 years; up-to-date]

For women who felt ambivalent about the current interval, less frequent screening was met with a more positive response and a recognition of the benefits. Perceived benefits included not needing as many screening appointments, the advantage of feeling reassured for longer and the potential to save the NHS money. For those who disliked the procedure, found it intrusive, worrying or inconvenient, the need for fewer screens was an obvious benefit of the interval change.

> *‘Especially as they’re not the most comfortable things to go through either. So that’d be a positive in the fact that you wouldn’t have to endure it every three years, or every two years… [*…*] you don’t have to go through it. The embarrassment.’* [P16; 30-39 years; up-to-date]

The new interval was also seen to provide reassurance for a longer period after a negative result. This view was more prominent among those without previous direct or indirect experience of abnormalities or cancer.

> *‘Um, and also, I guess that you know that you’re like clear for five years. I mean, in theory, because that would mean that the NHS is giving you the confidence that because they think it can be so long between screenings, you’ve had your screening and then you can have, like, the reassurance that, you know, I’m kind of good to go for five years.’* [P6; 25-29 years; up-to-date]
>
> *‘So, it’s kind of, it’s probably a positive, you know, that you’re not constantly being checked up on and made to feel guilty if it was meant to be more frequent and then you miss the frequency or, you know, things happen, if you go travelling, if you’re away for some time.’* [P6; 25-29 years; up-to-date]

Those preferring shorter screening intervals described being more likely to attend as soon as they were due, when a longer interval is implemented:

> *‘I think I’d make it more of a point to get there as soon as my screening was due. Just in case…I’d be getting myself there as soon as.’* [P5; 30-39 years; up-to-date]

However, some participants wondered if the new interval could have an impact on screening attendance, because a longer interval may make it easier to forget to attend or may reduce perceptions of urgency.

> *‘Maybe there’s even less of an impetus to respond to it and less of a sense of, it’s important to respond to it. So, I would be concerned that it kind of, the lengthening of the time somehow undermines its importance.’* [P13; 40-49 years; up-to-date]

Finally, for some, the potential for a new test to save the NHS money was a positive consequence of the change, as long as it also resulted in effective screening.

> *‘…if they can save money, then great. The main thing is, just making sure it’s effective. Yeah.’* [P10; 30-39 years; up-to-date]

### The process of accepting a longer interval

During the interviews, many of the women seemed to go through a process of increasing confidence in the interval change, moving from their existing acceptance of 3-yearly screening to an acceptance of 5-yearly screening. This was driven by their existing awareness of HPV primary screening and information provided during the interview to address initial concerns about the risks associated with a longer interval.

#### Awareness of HPV primary screening

Not all women were aware of the move to HPV primary screening, leading some participants to question why there was an interval change happening now. In the absence of an explanation, a range of potential reasons for the interval change were put forward, including the need to cut costs and the availability of HPV vaccination.

> *‘If it’s about like, capacity and saving money and I know it’s probably a big strain, you know, to do all of that, but if that’s the reason they want to make it a long period of time and this is the reasoning behind it, I think you’re dicing, you’re dicing with death a little bit there.’* [P5; 30-39 years; up-to-date]

Once HPV primary screening had been explained to them, some participants continued to question why a successful test such as the Pap smear would need to be changed at all.

> *‘I think there needs to be a valid reason as to why they’re changing it now. Why didn’t they start this from the beginning? [*…*] I don’t know why they did it every three years, but if they did it every three years, there must have been a reason why they did that.’* [P7; 30-39 years; up-to-date]

Additional questions about the implications of HPV primary testing were raised, including if the patient experience during the examination itself would be different and what would happen for people with abnormal cells or HPV-positive results (i.e., whether they would also be invited five-yearly).

> *‘So, will, the process, like when you’re actually in the nurse’s office change, or will that still be exactly the same?’* [P6; 25-29 years; up-to-date]

The information that screening would be more frequent for HPV-positive individuals was reassuring:

> *‘Knowing that if there is a problem in that HPV then they can check it out every year. Hopefully it will sort itself out. I think it’s actually better.’* [P22; 40-49 years; overdue]

### Understanding the rationale for the changes to a longer interval

Participants raised questions about the safety and effectiveness of the new interval and needed additional information to help build their confidence. For some, even the potential for a small risk would be a concern. These safety questions were raised particularly by women in their 20s to 30s.

> *‘But me personally, I would want to detect it as early as possible. Because you could be in that 1 or 2% like Jade Goody, you don’t know, there’s still a chance.’* [P8; 25-29 years; up-to-date]

Some women suggested that cervical cancer can appear between screens.

> *‘… you can get the HPV virus at any point in those five years. Right, so if you’ve had your test and you get it at year two, then you have to wait, you know, it’s like three years before you have your next test. And in that time, it could develop into cervical cancer. So, I’m wondering would it then be too late? I don’t know how quickly it develops and things like that.’* [P12; 30-39 years; up-to-date]

Concerns regarding the detection of HPV and cervical cancer also included the possibility of false negatives resulting from the use of the HPV test or the potential inability of the HPV test to identify abnormal cells without the presence of HPV.

> *‘So you-- So you could, very small chance, have abnormal cells without HPV, and that wouldn’t get picked up?’* [P4; 30-39 years; up-to-date]

These also included views that a longer interval might result in more adverse outcomes including more cancers, later diagnosis, more invasive treatment and more deaths:

> *‘Say there were abnormalities, but the test didn’t detect that, probably because they took very small amount of sample or whatever. And the test didn’t detect the abnormalities. And then there are five years now. So, in that-- She has to wait five years before she gets the next appointment, and then next appointment, they will discover that it has already happened, and the cells are already abnormal from the time the smear test was done when she was abnormal, but the test showed normal to five years, is it not going to be more risky for her life?’* [P14; 30-39 years; overdue]

Learning that the HPV test allows screening to pick up problems earlier in the cancer development process and that HPV infection takes a long time to develop into abnormal cells (more than five years), improved understanding of the interval change and was met with feelings of reassurance.

> *‘I feel much comfortable knowing the information I’ve got now. Especially when, you know, the time it takes to develop. Yeah, it makes complete sense. And yeah, I’d be totally comfortable with that.’* [P12; 30-39 years; up-to-date]
>
> *‘That makes sense to me. I would rather start at the HPV stage and know you’re being tested there rather than leaving it so it could possibly then develop onto the abnormal cells and then if it is every five years, from there I feel like it will probably be a lot better probably.’* [P15; 40-49 years; overdue]

### Influencing factors

Two factors seemed to influence acceptance of screening intervals: perceptions of personal risk (based on age and screening history) and feelings of trust and empowerment. For some women these factors meant that 5-yearly screening was still not acceptable, even towards the end of the interview process when messages about the rationale and safety had been presented.

#### Perceptions of personal risk

Participants reflected on their personal risk of developing abnormal cells and cervical cancer when describing their views about the existing and proposed intervals. Individuals with prior experience of abnormal results tended to feel that the new interval would pose a risk, even if small. They referred to their past results, their own safety, and worried that they might be ‘the rare case’ to develop aggressive, fast-growing cancer. Past experiences with cervical and other types of cancer in the family were mentioned.

> *‘I wouldn’t feel 100% comfortable. I really wouldn’t, but that’s because of my personal experiences. I think for anybody else, if it’s all clear and it’s always been clear then maybe, maybe it’d be alright. But I wouldn’t like that. I’d want that option of three years.’* [P19; 30-39 years; up-to-date]
>
> *‘…it then makes you worry even more because you think, hang on, does that increase my chances as well that it’s in the family and should I be really worrying here?’* [P21; 30-39 years; overdue]

Age was also seen to impact personal risk. Younger participants expressed the need for a more individual approach to screening (e.g. based on age) and felt they may need screening more frequently:

> *‘I’m just saying that I think it is a worry for young women and I think they should look at an age bracket and maybe test them a bit more often. Does that make sense?’* [P16; 30-39 years; up-to-date]

The reasons women gave for this included beliefs that younger women were at more risk of cervical cancer, that implications for future fertility were more important in women who had not yet had children and that younger women had generally had fewer screens.

> *‘Five years would just be too long. Yeah, because when you first start going for your smear test at age 25, then to leave it again until 30, that’s a big change. It’s a long time to leave. So yeah, in my opinion, I wouldn’t be happy with it being every five years.’* [P8; 25-29 years; up-to-date]

Older participants, by contrast, described themselves as feeling at lower risk:

> *‘No, I’ll be honest, I feel like because I am-- I never thought I’d say this, but because I am in the older bracket, I do feel a bit safer just knowing the causes of HPV and knowing that I’ve been fully checked out and things like that. So, I feel a lot more confident in terms of my having delayed my screening and not having gone in yet.’* [P15; 40-49 years; overdue]

Participants also assessed their risk based on their understanding of HPV transmission and their own relationship status, with those in longer term relationships and those who only had sex with women feeling at lower risk:

> *‘Yeah, I mean I suppose if you’ve not changed, you know, your partner or anything and you’re still in a relationship and you’ve not got the worst strain of HPV, I’m presuming that you would be happy to continue to do it every five years [*…*] but, you know, if there is someone that, you know, has got more sexual partners and they are, you know, more frequently and stuff like that, they would really not want to wait five years. Because it’s just like having a sexually transmitted infection, you might get treated for it and then go and pick it up again the next week.’* [P8; 25-29 years; up-to-date]

Perceptions of risk, especially in younger women aged 20-39 years, led some to raise the possibility of paying for private screening so they could be screened more often:

> *‘Could a woman, if they wanted to, sort of book in for a cervical smear more than the five-year period?’* [P1; 30-39 years; up-to-date]
>
> *‘…you’re forcing people to maybe go private for it and having to pay, and that’s a shame, because I would probably end up going down that route.’* [P3; 30-39 years; up-to-date]

Concerns were voiced regarding the ‘one size fits all’ approach to screening. A more personalised approach to cervical screening, taking into account a woman’s age and prior experiences (with HPV-positive results, abnormal cells, treatment, follow-up) was considered preferable.

#### Trust and empowerment

Trust and empowerment was also an overarching theme including perceptions of trust in healthcare professionals and decision-makers, and in science and research more generally. Some participants expected that any change to the screening programme would be a reasoned decision, based on consensus among experts. Trust in the government, the NHS, Public Health England and publicly funded bodies seemed to influence this:

> *‘…with the NHS within this country, I feel confident that they’re doing the right thing, so I’d be comfortable going along with what they think is right…I have faith that the public health are doing the right thing.’* [P16; 30-39 years; up-to-date]

Participants also expressed their trust in science and research findings and wanted to see more information about research evidence supporting the interval change:

> *‘Like, the government would need to put things out and be like, “This is what we’re backing. We know the science, and this is it, and this is okay,” to reassure a lot of women.’* [P3; 30-39 years; up-to-date]

For some participants, a change to extended intervals was seen as a cost-cutting exercise and sent a message about the value of women’s health. There was a particular tendency to express this view among those with previous experience of abnormal results.

> *‘-- and think, okay, well, maybe this, like, it could be based in science. It seems more likely to be, like, some kind of money-saving thing and then I would feel much less reassured.’* [P10; 30-39 years; up-to-date]
>
> *‘Well, these things are expensive, aren’t they? And then they’re trying to cut costs somewhere along the line and it’s usually women’s health, children’s health, old people’s health.’* [P22; 40-49 years; overdue]

Some described feeling powerless and frustrated about not being able to have more frequent screening, with a resignation that there was no choice but to accept the upcoming change.

This was expressed exclusively by women with previous experience of abnormal results (own or of family/friends).

> *‘Um, well, if they change it for five years, then you have no choice. One voice is not going to change it.’* [P14; 30-39 years; overdue]

## Discussion

This is the first study in England to explore, in depth, women’s attitudes to extending cervical screening intervals in the context of the move to HPV primary screening. Acceptability of extended intervals varied between participants and for many this changed over the course of the interview as additional information was provided. More than six months after the full roll-out of HPV primary screening across England, many of our participants were unaware of this change. Information about HPV primary screening and the rationale for the interval change led to a clearer understanding and, for some, acceptance of a 5-year interval.

Acceptability was influenced by views on the existing 3-year interval, which were underpinned by beliefs about the importance of prevention, the asymptomatic nature of cervical cell changes and perceptions of the timeline of cervical cancer development. It was also influenced by a range of factors including personal risk perceptions and trust in decision-makers.

Our study benefited from a sample that was diverse with respect to socio-economic status, ethnic background, relationship status and screening experience. However, it was also self-selected and it is likely that the views of women who are not interested in screening or in participating in research were under-represented. Although there were no explicit assumptions brought to the research, all researchers involved were interested in cancer prevention and LM/JW brought potential bias of past experience interviewing women about cervical screening and publishing on this topic. JB and MN had no previous experience in qualitative cervical screening research. We were able to identify positive as well as negative attitudes towards a change to the screening interval. The use of semi-structured interviews, during which women were provided with additional information, allowed us to explore how acceptability of extended screening might change as more information is provided.

Many of the themes raised towards the beginning of the interviews, prior to an explanation of the rationale for changing the interval, were similar to those highlighted in an Australian petition that followed implementation of extended intervals.(10) For example, women expressed preferences for more frequent screening, perceptions that cancer develops fast and beliefs that longer intervals would put lives at risk. De-valuing of women’s health and beliefs that the change was a cost-cutting exercise, were also expressed, in line with the Australian petition.(6) The views expressed following the provision of information were strikingly similar to another Australian study(11) which found that despite initially low awareness of HPV primary screening and understanding of HPV, concerns about longer intervals could be alleviated for many women by presenting information explaining the rationale for the changes, including the introduction of HPV primary screening, HPV-to-cancer development and the length of time this takes. These findings are also consistent with a quantitative study that showed women who were presented with this information had a better understanding of the change.(12)

Many of our findings could be mapped onto the Theoretical Framework of Acceptability (TFA).(13, 14) Of key importance was *intervention coherence*. Without a good understanding of the role of HPV in cervical cancer development, HPV primary screening and the rationale for the longer interval, women struggled to find the change acceptable. Many raised issues with the *perceived effectiveness* of the new approach, questioning the safety of the longer interval, and expressing concerns about false negative results and more adverse outcomes. *Affective attitude* appeared to be important in acceptance of existing intervals and extended ones. For some participants, there were positive attitudes associated with having to undergo an uncomfortable procedure less often (once they were reassured about safety). Participants also expected to feel positive emotions following a ‘normal’ result with a longer interval, which would be reassuring for a longer period. Others expressed worry and other negative emotions at the prospect of a change which they felt powerless to challenge. Our theme of ‘Trust and empowerment’ could be related to the TFA construct of *ethicality* and suggests trust in decision-makers will likely affect the extent to which the change is seen as ethical and therefore acceptable. *Opportunity costs, burden* and *self-efficacy* were less obviously important in this context.

Perceived risk was discussed throughout the interviews and was influenced by age, previous experience of abnormal screening results and family history. These seemed to impact acceptability of a longer interval, consistent with other work.(19) In a quantitative study exploring acceptability of interval changes following information exposure,^12^ participants who were classed as irregular screening attenders showed more positive attitudes towards the interval extension. We also found that individuals who were overdue for their 3-yearly screen tended to be less negative about the prolonged interval.

Our study suggests that communication about the rationale for extending screening intervals will need to include a clear explanation of current HPV screening. Some participants wanted to see an explanation of the scientific evidence supporting the interval change, including information on the sensitivity of the HPV test, timeline of abnormal cells and cervical cancer development, and robustness of studies on which the decision to extend intervals was based. Conveying the messages that HPV testing has the potential to intervene earlier in the oncogenic process than cytology, and that the timeline from HPV exposure to cancer development is long, is likely to help increase acceptability. Multi-level communication, including public awareness campaigns, print and online information, as well as conversations between trusted health professionals and those attending for screening will be needed to ensure widespread understanding and acceptance.

This study provided an indication of the information that is important for women to understand and accept the proposed screening interval change. However, more research, with larger samples, is needed to examine the impact of specific messages and identify the best way of communicating information about HPV testing and the rationale for longer intervals to screening-eligible women. In addition, it may be useful to identify demographic sub-groups who are less accepting of longer intervals so that targeted communication can be developed. Our findings suggested women who were regular screening attenders were more resistant to longer intervals but this needs further investigation.

## Conclusion

Women showed a range of cognitive and emotional responses to learning about a planned extension to cervical screening intervals. A number of factors influenced whether participants found the change to 5-yearly screening acceptable. These included attachment to the current 3-yearly interval, trust in decision-makers, and understanding of the rationale for the change. When women had the opportunity to discuss the changes and were provided with more information they were, for the most part, reassured and more accepting of the interval change. This highlights the importance of good communication.

## Supporting information

Appendix 1: Example quotes for each theme

## Data Availability

Data may be made available from the corresponding author upon reasonable request.

## Data Availability

Data may be made available from the corresponding author upon reasonable request.

## Contributorship Statement

JW and LM conceived the study. JB, JW and LM designed the study materials. LM and JB conducted the interviews. MN and LM analysed the data and drafted the paper. All authors contributed to the final version of the manuscript.

## Data availability

Data is available from the corresponding author upon reasonable request.

