## Appendix 1: Example quotes for each theme for "Acceptability of extending HPV-based cervical screening intervals from 3 to 5 years: An interview study with women in England": Supplementary File 1 - Appendix 1.pdf

| **Acceptance of existing screening intervals** |
| --- |
| *General acceptance* |
| "Ah, I suppose it's alright. I wouldn't mind if it was a little bit longer, but I suppose it's alright." [P11; 30-39 years; overdue] |
| "I think I'd rather go more often than not, if that makes sense? … So, yeah, three years seems like a decent amount of time to leave it." [P1; 30-39 years; up-to-date] |
| *Prevention and early detection* |
| "I think that prevention is really important, because if you can prevent or if you can catch something at the early stages, it's better than waiting." [P7; 30-39 years; up-to-date] |
| "If you can prevent it, do you know what I mean, it's one of the only cancers that you are able to prevent if you catch it early. You don't have routine tests for any other type of cancer really" [P8; 25-29 years; up-to-date] |
| "I don't actually know how many-- What the odds are of getting cervical cancer is. I have no idea of like, if leaving it too late, what would happen. So, all of that is, yeah. I guess it's lack of information that doesn't make you-- It's not making me move any faster, because I actually don't know, if I don't check it tomorrow, what's the outcome of that? Or what could be-- Yeah. I don't know." [P15; 40-49 years; overdue] |
| *Cervical cancer symptoms* |
| "If cancer of, you know, cervical cancer isn't caught early on when there's probably no symptoms, then it can be quite devastating. So, I think it's quite, it's very vital, really , that they have the tests, um , for people to try and pick up early. Yeah." [P4; 30-39 years; up-to-date] |
| "It feels like it's something that you can't be proactive about until it gets to a point where there's symptoms and presumably when there's symptoms, then some things have fairly significantly developed." [P13; 40-49 years, up-to-date] |
| *Timeline of cervical cancer development* |
| "When I think about cancer, I feel like it's something that's potentially done, it's quite quickly and can be life threatening..." [P13; 40-49 years, up-to-date] |
| **Acceptance of 5-yearly intervals** |
| *General acceptance* |
| "Um, so for me, if I had to wait five years, I guess I'd have it in the back of my mind of, oh, I wonder if everything is fine and normal." [P18; 30-39 years; up-to-date] |

“Oh my God, that's terrible. Awful. I'm shocked. 5 years is really bad...I feel like, I would really be unhappy if I had to wait every five years. That would make me feel quite ill. I'd be upset with that.' [P3; 30-39 years; up-to-date]

“That makes me quite nervous. That seems like quite a, quite a long time. That would mean, you know, between my first one and my next one, that would be from 25 to 30. I'd be 30 the next time I had one and that, to me, makes me quite nervous.' [P6; 25-29 years; up-to-date]

“Because once you tell some people they're gonna be like, ‘Hang on a minute. That's too long. Five years is far too long for me. I can't go without a screening.’ And if they're not told anything on top of that, then you can go away really worried and quite sort of scared about the idea of it.” [P20; 30-39 years; up-to-date]

*Contradicting existing beliefs*

“... if there's no symptoms until this extensive cell change. Then that feels like the five years I've got no recourse to any kind of medical support or help or intervention. Because I've got absolutely no idea what's going on.” [P13; 40-49 years; up-to-date]

“...it's just not something I would ever want to leave that long… you could go a few years and be okay and then all of a sudden you've got that strain of the virus again, and then that's developing into abnormal cells. And still, you've got another two years before the next test. So that could rapidly spread or, you know, rapidly turn more severe.” [P8; 25-29 years; up-to-date]

“Um, well, that sounds okay if you're testing earlier. But personally, I still think that five years is too long, even though you're testing earlier. It still could develop, couldn't it? So, I'd say five years is too long.” [P17; 40-49 years; up-to-date]

'Yeah, but then, in between-- Say there, say there were abnormalities, but the test didn't detect that, probably because they took very small amount of sample or whatever. And the test didn't detect the abnormalities. And then there are five years now. So, in that-- She has to wait five years before she gets the next appointment, and then next appointment, they will discover that it has already happened, and the cells are already abnormal. So, in a case like this, what are they going to do?” [P14; 30-39 years; overdue]

*Recognition of the benefits*

“... if you go, and then you know what you're looking for, the HPV, and then they will say, ‘Okay, no anomalies, you come back in five years,’ you're gonna trust that you actually have no problems. So, it is actually nice to know that I don't have to keep on going …" [P22; 40-49 years; overdue]

“The less you go, the better it is for an embarrassing-- From an embarrassment point of view…” [P5; 30-39 years; up-to-date]

“Right, so if they're normal [the cells] then it would be every five years instead of three. Okay. Yeah, I feel like it could, yeah, I guess it could make you feel quite reassured in the sense of, you know, your results are quite-- Well, they're normal, and so we're going every five years instead of three, could make some women feel as though that's like less pressure off them. And they're able to take that away from, you know, their health worries.” [P9; 25-29 years; up-to-date]

*Impact on screening attendance*

| |
|---|
| “I think-- As far as I can remember it used to be five years [the screening interval] and it's quite easy to forget after that length of time to actually go to your appointment. I know they send you letters and stuff, but people move, don't they and what have you? So, yeah, it can be-- If it's more frequent then you're more likely to remember to go.” [P1; 30-39 years; up-to-date]<br><br>“I think it [the interval increase] would just be, it would just increase my, not awareness of it, but increase my thinking, you know, I'd be focusing more on that. You know, every three years seems to be just nice at the moment. But any more than that and I'd be thinking, you know, I've got another year until I have my smear test and things like that.” [P12; 30-39 years; up-to-date] |
| **The process of accepting a longer interval** |
| *Awareness of HPV primary screening*<br><br>“If the current plan is working, in terms of its reducing the incidents of cancer or deaths from cancer or whatever, why would we be changing it?” [P13; 40-49 years; up-to-date]<br><br>“Right, so it's switching to five years for the HPV. But will the smears still be getting done as well or not? … Yeah, I mean, I think when you first said that I thought you meant they were going to be doing the HPV test alongside the smear.” [P8; 25-29 years; up-to-date]<br><br>“Okay, um, another thing-- Is this because a majority of the female population have had their HPV vaccine, so is that why they want to, is that why they think that if they've had a vaccine they will, you know, have a positive result with HPV? Which means that if they're already vaccinated and they have their cervical screening, of course, there'll be less risk of them developing abnormal cells, that's why they want to move it to five years?” [P7; 30-39 years; up-to-date]<br><br>“Okay. But would the test be exactly, would the test be exactly the same like they do currently? I will want to know whether the test they are going to do, the fact that it's changing, would it be slightly different or would it be the same? Or will it take longer for them to give me a response or if it is abnormal, what would be their steps?” [P14; 30-39 years; overdue]<br><br>*Understanding the rationale*<br><br>“I probably-- I think I'd need information on how-- So if you get the HPV, if you get that virus , how long it takes for that to turn into abnormal cells or potentially, I'd need to know the time frames to figure out whether I think five years is a good time frame. Because obviously, I said before the three years I thought was too long because something could happen in the first few months after a smear test and then you've got to wait years. But it could actually take years for the virus to turn into abnormal cells. So, I don't know the time frames.' [P4; 30-39 years; up-to-date]<br><br>“So probably like I've mentioned, how quickly the stages progress and if that time period is, like, safe to be able to detect something from, like, you know, incubation until, like, actually getting cancer.' [P6; 25-29 years; up-to-date]<br><br>“The main thing is, just making sure it's effective. Yeah.' [P10; 30-39 years; up-to-date]<br><br>“Yeah, so-- So people that have abnormal cells, do they all have HPV, I assume?' [P4; 30-39 years; up-to-date] |

“I'd want to be sure. I'd want to know. I want to know a bit more about, like, why they think that it's sooner...I want to know the theories behind why they think that five years is acceptable with obviously the testing.' [P3; 30-39 years; up-to-date]

“Yes, I mean if they manage to catch it in the earlier stage, that would be really good, yeah.' [P11; 30-39 years; overdue]

“I feel much comfortable knowing the information I've got now. Especially when, you know, the time it takes to develop. Yeah, it makes complete sense. And yeah, I'd be totally comfortable with that.” [P12; 30-39 years; up-to-date]

“Oh okay. I mean, when you-- When I have the kind of like time frame, I feel much better about it, you know? And I think that's probably like what's missing from a lot of, yeah, the way I'm thinking about it and also, it seems like from what you're saying that the type of cancer or the type of cells that you're kind of screening for in a smear, you know, it's very different from other types of cancer. And I guess, like, in my head, all types of cancer get kind of lumped into one type, and I guess that's where a lot of the misunderstandings happen. I think that's quite reassuring. I'd still look a little bit suspicious but, yeah.” [P10; 30-39 years; up-to-date]

“...at the end of the day , if there's a way for them to directly test for the HPV and, you know, you've got the stage one , then I can understand why would be every five years , especially if your results come back as normal , then yes , that would make sense. Yeah.”[P9; 25-29 years; up-to-date]

“That kind of explains quite a bit, actually. Which I didn't even know. So that kind of makes things a lot more clear around the whole process of it and obviously, there is a step before... Because if you're told about from the beginning and the abnormalities stage and then later down the line, if you do develop cervical cancer. So, to know about that as well, that's very useful. And obviously understanding the process now, why they do have longer in between screenings, is because they are testing for the infection first so that can eradicate it later down the line. So that's really useful to know.” [P20; 30-39 years; up-to-date]

“And I suppose I can't get away from this idea that if you have a screening, it's-- And you get a clear result, it's clear at that point. What is it that then happens that makes cells start to change? And what happens-- I don't know. What happens then between that point and five years later, or four and a half years, or four years later? What happens to those cells?” [P13; 40-49 years; up-to-date]

**Influencing factors**

*Perceptions of personal risk*

“Um, I don't know, because I had these noncancerous cells which I had removed, if I was given that option of, "Okay, everything's clear, you're all alright, we could see you in the next four or five years," I'd still ask them, you know, "can I still be seen in three years?" ' [P19; 30-39 years; up-to-date]

'” think I'd feel a bit more-- I'd feel a bit nervous about that, just for the sake that, I mean, because I've had the experience of having the cell abnormalities sort of thing. You know, if I had been fine all throughout then it'd be fine. I wouldn't think twice about it. But yeah, I wouldn't want it to be any more than three years.” [P12; 30-39 years; up-to-date]

| |
|---|
| “And um, you know, I'm married, so obviously, if I didn't have the HPV and then I went for my test and they say, "Well you still haven't got it", well, the chances are, you know, unless I do something naughty that I'm probably not going to get it. Is that-- Am I thinking that right?” [P1; 30-39 years; up-to-date]<br><br>“Um, I guess it seems like a bit too long, but that opinion might be coloured by the fact that my mum did have to have a hysterectomy because she had a lot of pre-cancerous cells” [P18; 30-39 years; up-to-date]<br><br>*Age as a risk factor*<br><br>“That means if I want one, I have to go private, but I shouldn't have to go private for something so, you know, normal and so, you know, regular.” [P2; 25-29 years; up-to-date]<br><br>“Um, I don't know, would there be like a procedure in place where women would be able to request having it earlier? Or would that just be the standard every five years, maybe?” [P9; 25-29 years; up-to-date]<br><br>*A personalised approach*<br><br>“And I think, I don't-- What I'm saying is I don't think everyone should be bucketed into those three years. I think if you're over a certain age maybe, then get it more often. If you are, um, in a bucket, there's higher risk. Then you should be getting it more often. I don't think it's fair to say, ‘Everyone's either three years or every year.’ That just doesn't really work... So, it just, as if, you know, with an assumption that a 20-- A healthy 27, 28-year-old female. You know, it just, I don't know, it just seems silly to not, to wait five years.” [P2; 25-29 years; up-to-date] |
| *Trust and empowerment*<br><br>“Our health service is in such an almighty state that-- I mean, people tell me all sorts of absolute nonsense in the past, that I just kind of think, well, I'm not, I'm just not convinced by it.” [P13; 40-49 years; up-to-date]<br><br>“I know we have a very, very good health care system despite what gets said in the media sometimes. But I, I suppose I trust that we do it [screening] for three years for a reason.” [P21; 30-39 years; overdue]<br><br>“Um, yeah, more information sent to us would be ideal. I'd also like to see, um, in a few years obviously, statistics from 2019. Um, obviously they'd need, I don't know, 5, 10 years to see if it has actually changed anything.” [P5; 30-39 years; up-to-date]<br><br>*The value of women’s health*<br><br>“I think in general and sort in medical science, um, it does feel like women are sort of getting like, well, maybe not so much anymore, but it sort of feels that women can get left behind a little bit.” [P1; 30-39 years; up-to-date]<br><br>“It doesn't sound-- It doesn't sound like they're taking into consideration the health of people; it sounds more like they're putting costs first.” [P7; 30-39 years; up-to-date]<br><br>*Feeling powerless and frustrated* |

“I still-- I mean, if a decision was taken to every five years, then you'd have to accept it…” [P21; 30-39 years; overdue]

“I think three years is kind of too long, so if they made it even longer. Yeah. I don't think I'd be, you know, I'd probably have to deal with it…” [P4; 30-39 years; up-to-date]
